# Application of a generalized SEIR model for covid-19 in Algeria

**DOI:** 10.1101/2020.08.10.20172155

**Authors:** Mohamed Lounis, Juarez dos Santos Azevedo

**Affiliations:** Department of Agro-veterinary Science, Faculty of Natural and Life Sciences, University of Ziane Achour, BP 3117, Road of Moudjbara, Djelfa 17000, Algeria.; Universidade Federal da Bahia (UFBA), Instituto de Ciências, Tecnologia e Inovação, Centro,42802-721, Camaçari-BA, Brazil

**Keywords:** Epidemic Model, Covid-19, SEIR model, SEIRDP model, Algeria

## Abstract

The novel coronavirus diseases 2019 (COVID-19) in Wuhan is continuing to impress the world by its fast spread and the number of affected persons attracting an unprecedented attention. In this article, we used the classical SEIR model and a generalized SEIR model called SEIRDP model inspired in a model previously used during the outbreak in China to predict the evolution of COVID-19 in Algeria for a future period of 100 days using official reported data from early April to early August, 2020. Initial evaluation showed that thetwo models had a net correspondence with the reported data during this period for cumulative infected cases but the number of cumulative deaths was underestimated with the classical SEIR model. Model prediction with the SEIRDP concluded that the number of cumulative infected cases will increase in the next days reaching a number of about 60 k in middle November with a median of about 300 daily cases. Also, the number of estimated deaths will be around 2k. These results suggest that the COVID-19 is ongoing to infect more persons which may push national authorities to carefully act in the probable leaving of containment.

## Introduction

The coronavirus diseases 2019 (COVID-19) detected first in Wuhan, in the province of Hubei (China) on December 2019 has provoked a real confusion in the world [1]. This respiratory disease caused by a novel virus called SARS-CoV2 (Severe Acute Respiratory Syndrom coronavirus 2), has afflicted more than 19.4 millions positive cases and more than 723k deaths around the world [2].

Since the first case reported on February 25^th^, Algeria accounts currently 34,693cases and 1,293 deaths [3].

Due to the early and the drastic implemented measures in Algeria, the situation seemed to be under control until the last April and the beginning of May [1]. However, the evolution of the epidemic in the last two months has shown a real increasing in the daily reported cases. This may be related to the increasing in the total number of test performed but also to a result of the alleviation of the prevention strategy and the subsequent lightening in the respect of physical distancing and protection measures[1, 4]. In this way, comprehension of the epidemic curves and forecasting its evolution is very important for the evaluation of the implemented measures and in deciding to adopt the best future strategies to curb the spreading of the disease. In this way, statistics or mathematical modeling is of great importance and represents a crucial tool in understanding the epidemic characteristics and predicting its curve [5].

Multiple models have been achieved in Algeria to forecast the epidemic or to evaluate the preventive implemented measure with different approaches including local, regional and global studies [4, 6, 7,8, 9, 10, 11, 12, 13, 14, 15, 16, 17, 18]. These studies used the most common SIR[6, 7] and SEIR [4, 8, 9, 15,19] models or other mathematical models such as AFRIMA (Autoregressive Fractionally Integrated Moving Average Models) [12]and the Alg-COVID-19 Model [14].

In the current study, we used the classical model and a generalized SEIR model called SEIRDP (Susceptible, Exposed, Infected, Recovered, Death and Insusceptible (P) to predict the evolution of the epidemic curves in Algeria for a future period of 100 days.

### Data sources

The daily COVID-19 new cases (confirmed with RT-PCR test) in Algeria were collected from the Ministry of Population Health and Hospital Reform official website [3] from April 1^st^, 2020 to August 4^th^, 2020 for a time series of 126 days.

The file was used to build a time-series database including for each source the time series related to the total positive cases, the daily positive cases, the total deaths, the new daily deaths, the total recovered, the daily recovered and at last the number of quarantined persons. The total case in the mentioned period is of 31788 cases with an average of about 252 cases per day.

## Methods

The classical Susceptible Exposed Infectious Recovered (SEIR) model represents one of the most adopted mathematical models for characterizing and forecasting the epidemic diseases [4, 5]. It is widely used to study the COVID-19 curve in multiple countries in the world [4]. In the current study, we followed the adapted SEIR model developed by Tan and Chen [5], the most reliable response to emergency public health actions which takes into account the population exposed to the virus.

More precisely, we used the generalized version of the classical SEIR called SEIRDP (Susceptible, Exposed, Infected, Recovered, Death, Insusceptible (P)) a particular case of the model proposed by [19]. (see Fig. 1)

**Figure 1:**
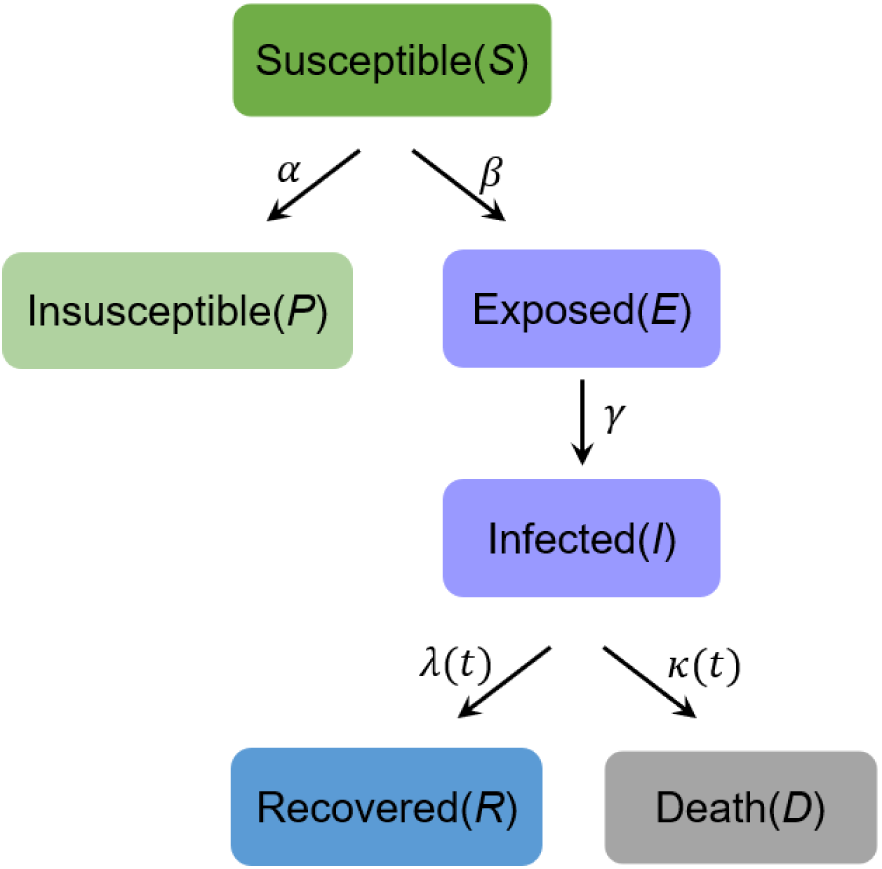
SEIRDP model scheme

In this model it is possible to include key epidemic parameters for COVID-19, such as the latent time, infected time, protection rateand time evolution of the recovery. This allows us to estimate the inflection point, ending time and total infected casesin Algeria according to the Ministry of Health of Algeria data.

### SEIRDP model

The SEIRDP (Susceptible, Exposed, Infected, Recovered, Death, Insusceptible (P)) model used to analyze the coronavirus epidemic in Algeria has the following variables:

- *S(t)*: Susceptible population;
- *E(t):* Population who are exposed to the virus, but not yet infectious in latent period;
- *I(t):* Population who get laboratory positive confirmation and with infectious capacity;
- *R(t):* Recovery cases;
- *D(t):* Death number;
- *P(t)*: Insusceptible cases;
- *N = S + P + E + I + R + D* is the total population;
- *α*: Protection rate (include people exposed to the infectious patients and people exposed to the asymptomaticpatients);
- *β*: Infection rate (include exposed people catch COVID-19, people get COVID-19 with diagnosed confirmationand people; without symptoms but can transmit it to others);
- *γ*-1: Average latent time;
- *λ(t)*: Coefficient used in the time-dependent cure rate;
- *κ*(t): Coefficient used in the time-dependent mortality rate.

The variables described above are related through the following ODE system:

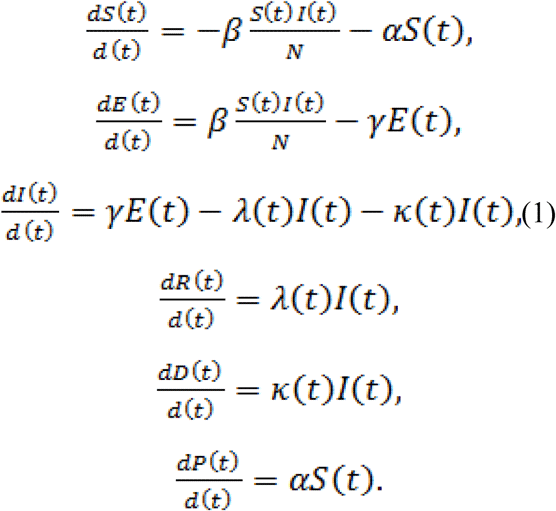

The model is time-dependent, because the cure rate λ(t) and mortality rate *κ*(t) are analytical functions of the timedefined by:

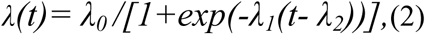

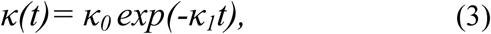

respectively, which are which are fitted according to the number of cases for *t* ϵ [*t*_0_; *t_f_*],where *t*_0_ and *t_f_* are the initial andfinal periods, respectively. Equations (2) and (3) indicate that cure rate or recovery rate *λ(t)* goes up exponentiallytoward a threshold value and the death rate κ(*t*) should be zero after an infinite time. Moreover, the parameters{*α*, *β*, *γ*^-1^, *λ(t)*, *κ(t)*} were fitted in the least square sense. In Figure 2 we plot these functions considering initial guess [λ_0_;λ_1_; λ_2_] = [0:1; 0:1; 0:1] and [κ_0_; κ_1_] = [0:5; 0:3] respectively.

**Figure 2:**
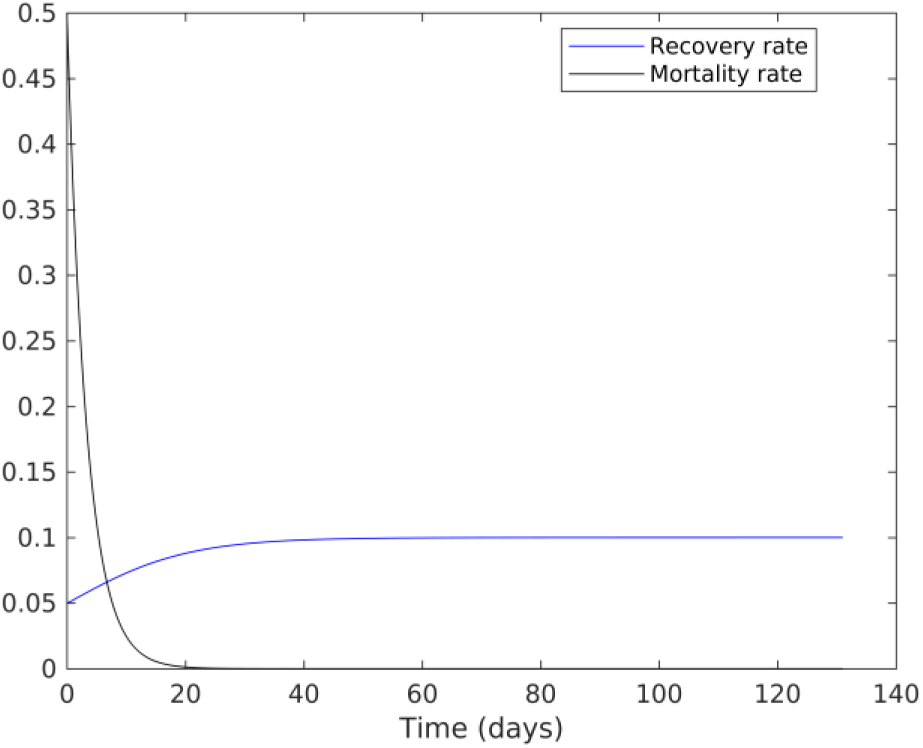
Plot of recovery and mortality rates.

The parameters {*α*,*β*, *γ ^-^*^1^,λ_0_, λ_1_, λ_2_, *κ*_0_ *(t)*, *κ*_1_*(t)*}are computed simultaneously by a nonlinear least-squares solver [20].

We can replace them in the model (1) and calculate the time-histories of the different states {*S(t);P(t); E(t);I(t);R(t);D(t)*}. In this case, we use the standard fourth–order Runge–Kutta process [21]. For this, the system of equationsis written as a single ODE such as:

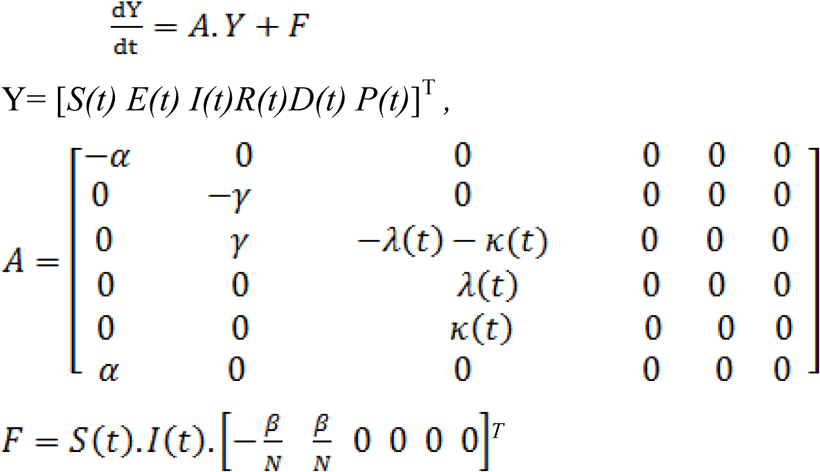

## Results and discussion

### Prediction of time evolution of COVID-19 for Algeria

Here we present the time series obtained by the SEIRDP model. The prediction of the Algerian situation according to this model shows the trends given in Fig. 3.

**Figure 3:**
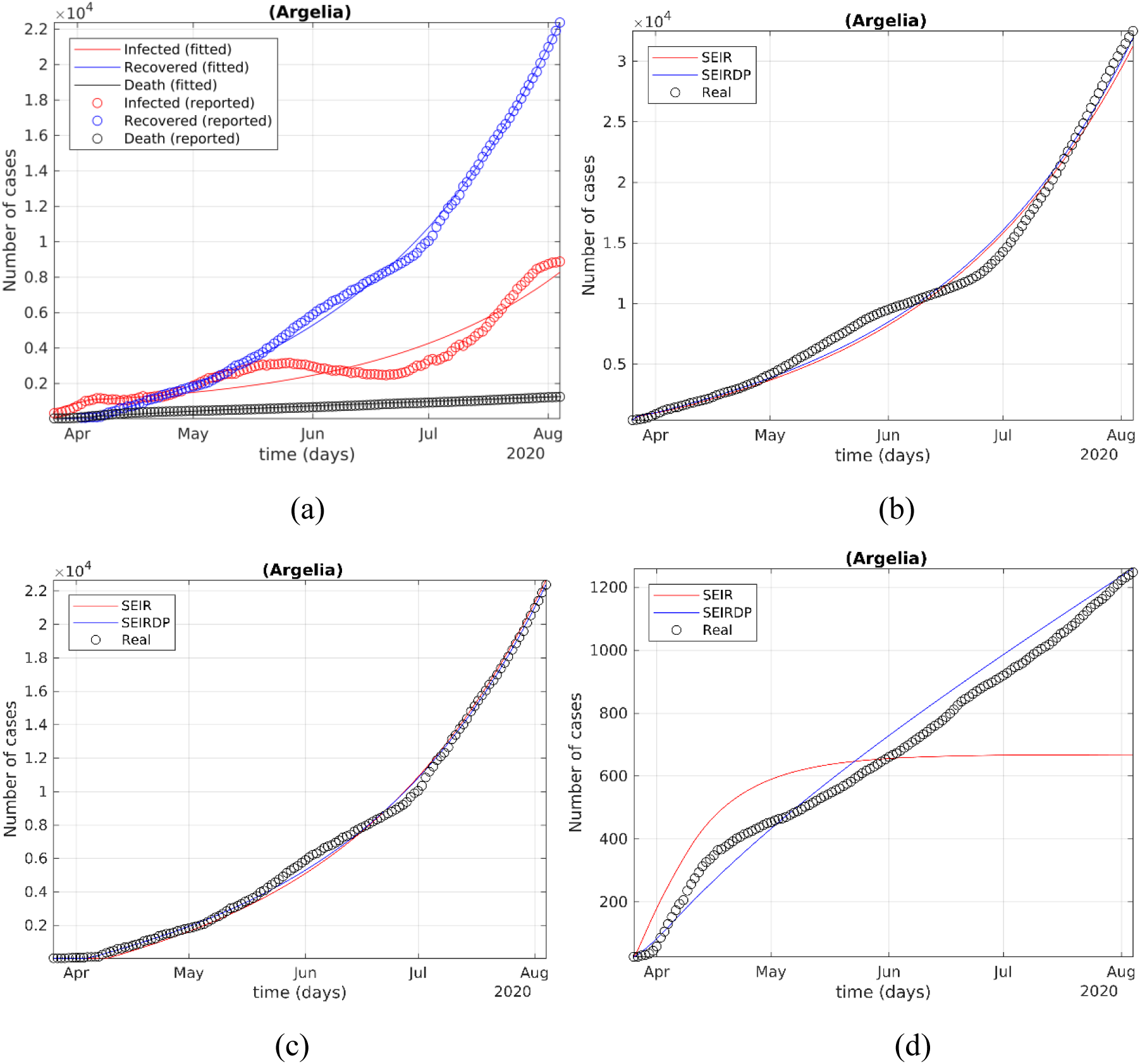
(a) Modeling of the differential SEIRDP model from April 1 to August 3. (c) Modeling of the differential SEIR model from April 1 to August 3 for cumulative positive cases (b), recovered(c) cumulative deaths (d).

The prediction of this model from early April, 2020 to early august, 2020 shows that our model has a good correspondence with the data reported in the same period by the ministry of health [3] in term of total, infected, recovery and death cases (Fig. 3(a)).This suggests that the state parameters found by the least-squares solution should be suitable for projecting future dates.

When compared with the traditional SEIR, results showed that prediction of the SEIR model was in correspondence with the prediction of SEIRDP and the reported cumulative cases (see Fig. 3(b)) and recovered cases (see Fig. 3(b)) by the ministry of health, while the number of predicted deaths was underestimated (see Fig. 3(d)).

We used later the SEIRDP model to predict the evolution of the pandemic for from April to November, 2020. Results showed that the number of cases in Algeria still increasing to attempt a number of about 60k positive cases and about 2k deaths in 12^th^, November, 2020 (see Fig 4(a). Prediction of the classical SEIR model however, estimates the number of cumulative cases at about 46k (see Fig 4(b). In Figs. 4(c)-4(d) we have the recovery and death rates as a function of time respectively. In this case, we compare the results between the computed and observed rates.

**Figure 4:**
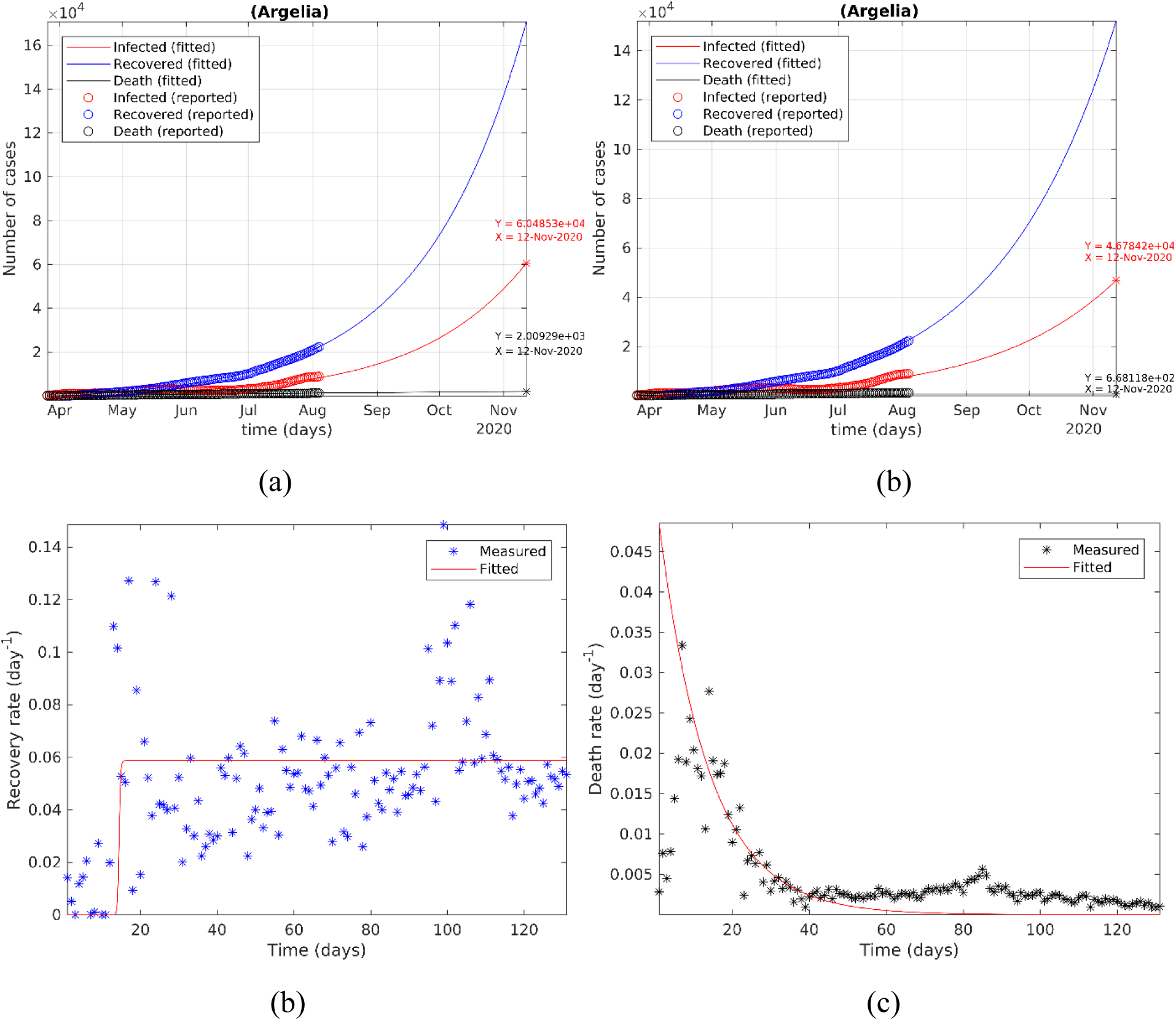
(a) Predictions of the differential SEIRDP model for Algeria from early April to mid-November, 2020. (b) The recovery rates and (c) deceased rates of April to early-August, 2020.

In Fig. 4(b) we can observe a constant behavior while in Fig. 4(c) we have an exponential decrease in mortality rates as time progresses.

These results corroborate with the plots of Fig. 2.

Of notes, the current model is using the official data by the Ministry of healthy based only of RT-PCR testing. The number of performed tests was very low in the first days (about 60 tests/day). This number has currently attempt 2500 tests/day but remain low for screening the total cases witch could affect the total number of cases in the model.

## Conclusion

In this study, we used a classical and a generalized SEIR models to analyze the epidemic curve COVID-19 and its evolution in Algeria.

Based on data reports of the Ministry of public health and hospital reform from early April to early August, we first showed that the two models, the classical SEIR model and the SEIRDP model had a good correspondence with the daily reported data in term of cumulative infected cases and recovered while the classical SEIR model underestimated the cumulative deaths. The SEIRDP model prediction for the next 100 days showed that the number of cases will increase to reach a number of 60k in middle November while the predicted number of deceased persons will be around 2k cases. The number of cumulative cases with the classical SEIR model will reach a number of about 46k in the middle November. These results showed that the number of cases is still increasing in Algeria despite the implemented measures which may be taken in consideration public authority in the next steps of fighting this disease.

## Data Availability

have followed all appropriate research reporting guidelines and uploaded the relevant EQUATOR Network research reporting checklist(s) and other pertinent material as supplementary files, if applicable.

## Notes

### Competing Interest Statement

The authors have declared no competing interest.

### Funding Statement

Not concerned

